# Patient Perceptions of Nursing Care Quality: Insights from a Public Hospital in Mexico

**DOI:** 10.1101/2025.07.17.25331752

**Authors:** N.P. Muñiz Ojeda, F.J. Frausto Quiroz, David A. Franco-Torres, Agustín Avalos, Mauricio Sánchez-Barajas

## Abstract

Patient-centered care is a cornerstone of healthcare quality, with nursing professionals playing a pivotal role in addressing patients’ needs through effective interpersonal relationships. This cross-sectional observational study assessed the perceived quality of nursing care among 374 hospitalized patients at the General Hospital of Zone / Family Medicine No. 21 in León, Guanajuato, Mexico. Data were collected using the validated Patient-Perceived Nursing Care Quality Evaluation Questionnaire (CECOP), focusing on empathy and respect within the nurse–patient relationship. Descriptive statistics were employed to characterize the sample and summarize patient perceptions. Results indicated that 47% of patients rated nursing care as regular, 42% as good, and 11% as bad. Notably, patients with lower educational attainment more frequently perceived care as regular, whereas those aged 60–70 years predominantly rated care as good. These findings highlight the need to enhance communication and humanized nursing practices through continuous professional development to improve patient satisfaction and health outcomes.

## 1 Introduction

The Pan American Health Organization (PAHO) defines quality of care as a people-centered process that emphasizes the needs of individuals, families, and communities, and is characterized by attributes such as safety, effectiveness, timeliness, efficiency, and equitable access. Attaining these standards requires well-organized, adequately resourced, and humanistically guided healthcare services. Within this framework, nursing professionals—who maintain direct, continuous contact with patients—play a critical role in determining care quality, particularly through their ability to communicate effectively, demonstrate empathy, and uphold patient dignity.

Nursing care quality is often assessed through the lens of patient satisfaction, which reflects the alignment between patient expectations and their actual experiences. When this alignment is absent, dissatisfaction may arise, revealing the perceived shortcomings of care delivery. Among the key elements influencing this perception is the interpersonal relationship established between nurses and patients, which encompasses both technical competence and relational behavior. As such, communication, respect, and emotional support are increasingly recognized as central to humanized, high-quality nursing care.

Contemporary healthcare systems, however, face operational pressures—limited time, high patient volumes, and administrative demands—that risk reducing nursing interventions to mechanized tasks, thereby neglecting the relational components of care most valued by patients. This tension underscores the importance of evaluating care not only through clinical outcomes but also through the subjective experiences of those receiving it.

In line with this approach, theoretical models such as Jean Watson’s Theory of Human Caring and Avedis Donabedian’s framework for quality assessment emphasize the moral and affective dimensions of care. These perspectives advocate for a holistic, compassionate practice that fosters trust, respect, and well-being, ultimately improving patient outcomes and satisfaction.

The present study investigates hospitalized patients’ perceptions of nursing care quality, with particular attention to the interpersonal dimension of nurse–patient interactions. Using the validated CECOP instrument (Patient-Perceived Nursing Care Quality Evaluation Questionnaire), which focuses on communicative behaviors such as empathy and respect, the study aims to provide insight into how patients experience the quality of nursing care in a high-demand institutional setting. Conducted at the General Hospital of Zone and Family Medicine No. 21 in León, Guanajuato—part of the Mexican Social Security Institute (IMSS)—this research contributes to broader efforts to humanize hospital care and strengthen the professional development of nursing.

## 2 Methods

An observational, cross-sectional study was conducted with a total of 374 hospitalized patients of both sexes at the General Hospital of Zone / Family Medicine No. 21 in León, Guanajuato, Mexico. Participants were selected through a non-probabilistic convenience sampling approach and included individuals from various hospitalization wards to ensure diversity within the sample.

Data collection involved the use of an item derived from the *Patient-Perceived Nursing Care Quality Evaluation Questionnaire* (CECOP), a validated instrument designed to assess patients’ subjective evaluations of nursing care. For the purposes of this study, only the global perception item was employed, which asked patients to classify the overall quality of nursing care received as good, regular, or bad. This approach focused specifically on the patient’s general impression of care, emphasizing the interpersonal aspect of the nurse–patient relationship.

Sociodemographic data, including age group and educational attainment, were also collected to explore trends in perception across different patient profiles. Descriptive statistics—frequencies, percentages, means, and standard deviations—were used to summarize the sample characteristics and distribution of responses. Stacked bar graphs were generated to visually represent the levels of perceived nursing care quality according to educational level and age group.

All data were organized and processed using Microsoft Excel. Given the descriptive scope of the analysis, no inferential statistical tests were applied. The study received ethical approval from the Institutional Review Board and the local Ethics Committee. Informed written consent was obtained from all participants prior to data collection. The research adhered to the ethical standards established in the Declaration of Helsinki and the Belmont Report, ensuring respect for participant autonomy, confidentiality, and well-being. A sample size of 374 patients was maintained to ensure adequate representativeness and minimize data loss.

## 3 Results

A total of 374 patients completed the survey assessing the quality of nursing care. The overall distribution of responses indicated that 11% of patients rated the nursing care as “bad,” 42% as “good,” and 47% as “regular.” Further stratification by educational attainment revealed that patients with only primary education were more likely to perceive the quality of care as “regular.” Conversely, age-based analysis showed that patients aged 60 to 70 predominantly rated the nursing care as “good.” These results highlight variations in perceived nursing care quality influenced by patients’ educational level and age group.

### Percentage Distribution of Patient-Perceived Nursing Care Quality

**Figure 1:**
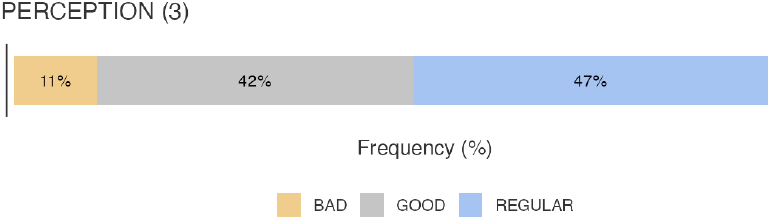
Proportion (%) of total patients who rated nursing care as good, regular, or bad.

The frequency distribution of patient responses showed that 41 participants rated the quality of nursing care as “bad,” 157 as “good,” and 176 as “regular.” This distribution indicates that nearly half of the respondents perceive the care as moderate (“regular”), while a significant proportion consider it satisfactory (“good”).

The relatively smaller group rating the care as “bad” suggests that while there are concerns about nursing care quality among some patients, the majority experience a level of care ranging from moderate to positive. These frequency patterns underscore the importance of targeted quality improvement initiatives to address the needs of patients expressing dissatisfaction.

### Frequencies of Nursing Care Ratings

**Figure 2:**
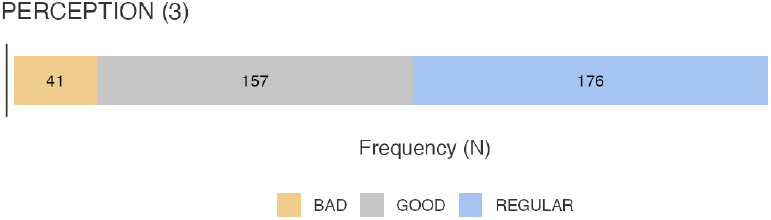
Frequency Distribution of Nursing Care Ratings.

**Figure 3:**
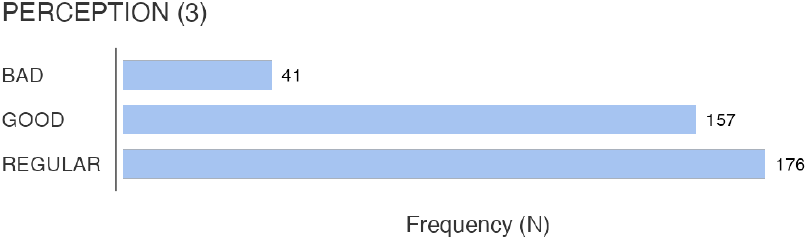
Frequencies (n) of patients who rated care as good, regular, or bad.

Among the 374 patients surveyed, perception of nursing care quality varied across different educational levels. Patients with only a primary education most frequently rated care as “regular” (n = 75), followed by “good” (n = 62), and “bad” (n = 10). Those with secondary education similarly rated care as “regular” (n = 61) and “good” (n = 56), though the number of “bad” ratings was higher in this group (n = 18).

Among patients with a high school education, 29 perceived the care as “regular,” 26 as “good,” and 6 as “bad.” Finally, patients holding a bachelors degree reported 14 “regular,” 12 “good,” and 5 “bad” ratings. These findings suggest a consistent trend in which patients with lower educational levels tend to perceive nursing care as “regular,” while those with higher education levels provide a more balanced distribution of ratings, including relatively fewer “bad” assessments.

### Detailed Frequency of Nursing Care Ratings by Educational Attainment

**Figure 4:**
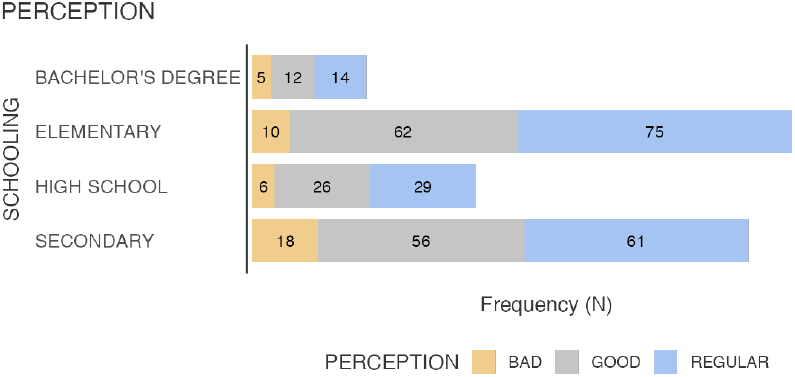
Number of patients in each educational group rating nursing care as good, regular, or bad.

The perception of nursing care quality varied across different age groups among the surveyed patients.

In the 20–29 years cohort, 33 participants rated care as “good,” 25 as “regular,” and 11 as “bad.” Patients aged 30–39 predominantly perceived care as “regular” (n = 40), followed by “good” (n = 28) and “bad” (n = 14). Similarly, those in the 40–49 age group rated care mostly as “regular” (n = 39), with fewer “good” (n = 18) and “bad” (n = 7) ratings. Among patients aged 50–59, the majority rated care as “regular” (n = 48), while 27 considered it “good,” and 4 rated it “bad.”

In contrast, patients aged 60–70 predominantly rated nursing care as “good” (n = 49), with fewer perceiving it as “regular” (n = 27) or “bad” (n = 4). Overall, despite a substantial proportion of patients across all age groups rating nursing care as “good,” the prevailing perception among most respondents was that of “regular” care quality.

### Frequency Distribution of Nursing Care Ratings by Age Group

**Figure 5:**
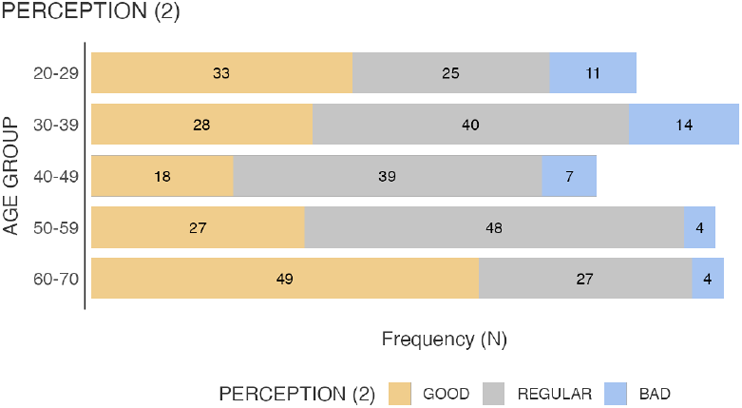
Number of patients in each age cohort who rated nursing care as good, regular, or bad.

## 4 Discussion

Previous studies have reported varied levels of patient satisfaction regarding nursing care quality. Ruiz-Cerino et al. found that approximately two-thirds of participants were satisfied, with one-fifth highly satisfied with nursing care received. Similarly, Burgueño et al. observed that the majority of patients rated nursing care as excellent (69.4%), followed by very good (13.3%) and regular (1%), indicating a predominantly excellent perception of care quality. Velarde del Río reported that 67.9% of patients felt highly satisfied with the nursing care, while 27.7% expressed moderate satisfaction.

Rodríguez et al. emphasized that nursing staff must deliver care grounded in principles and values, ensuring timely, continuous, and respectful attention to patient integrity. Becerra et al. identified that around one-third of patients perceived intermediate, low, and high levels of care quality equally, highlighting the critical need for ongoing training and professional development to guarantee quality, person-centered care.

Another study reflected results consistent with the present investigation, where 40% of participants rated care quality as regular, 37.1% as good, and 22.9% as deficient. These findings align with the current study, suggesting that nursing care quality is generally perceived as ranging from good to regular by most patients.

## 5 Conclusion

This study examined hospitalized patients’ perceptions of nursing care quality at the General Hospital of Zone / Family Medicine No. 21 in León, Guanajuato, Mexico. The results reveal that the majority of patients rated the overall quality of nursing care as either “regular” (47%) or “good” (42%), with a smaller proportion (11%) reporting it as “bad.” These findings indicate that while a significant number of patients recognize satisfactory care, a notable segment perceives the quality as only moderate, highlighting room for improvement in the nurse–patient relationship.

Educational level and age appeared to influence patient perceptions. Patients with lower educational attainment tended to rate care as “regular,” whereas those aged 60 to 70 years were more likely to rate it as “good.” This suggests that sociodemographic factors may shape patients’ expectations and evaluations of nursing care.

Given that the interpersonal dimension—particularly respect, empathy, and communication—plays a pivotal role in patient satisfaction, the findings underscore the importance of reinforcing humanized care practices. Investment in continuous professional development, with an emphasis on communication skills, emotional support, and respectful interactions, is essential to elevate the quality of care perceived by patients.

Ultimately, improving these relational aspects of nursing care will help foster trust, enhance patient experiences, and contribute to more compassionate, effective, and patient-centered hospital environments.

## Acknowledgments

The authors extend their sincere gratitude to the nursing and administrative staff of the General Hospital of Zone and Family Medicine Clinic No. 21 (HGZMF No. 21) for their invaluable support during participant recruitment and data collection. Special thanks are also due to the Department of Medical Education and Research for their assistance with data organization and visualization. Above all, the authors are deeply grateful to the patients who generously shared their time and experiences, making this study possible.

## Authors’ Contributions

All authors contributed equally to the conceptualization, design, data collection, analysis, and writing of the manuscript. Each author reviewed and approved the final version of the manuscript and agrees to be accountable for all aspects of the work.

## Data Availability Statement

The datasets generated and analyzed during this study are not publicly available due to institutional privacy regulations. However, data may be made available upon reasonable request to the corresponding author, subject to approval by the Instituto Mexicano del Seguro Social (IMSS) and in accordance with institutional data-sharing policies.

## Ethical Approval

This study was approved by the Research Ethics Committee of the Mexican Social Security Institute (IMSS) and conducted in accordance with the principles of the Declaration of Helsinki and national regulations (NOM-012-SSA3-2012). Written informed consent was obtained from all participants prior to enrollment.

## Declaration of AI Usage

No artificial intelligence tools were used in the conception, drafting, data analysis, or writing of this manuscript. All scientific content, language editing, and interpretation of findings reflect the authors’ own intellectual work. The manuscript was typeset using LaTeX via the Overleaf platform.

